# The Impact of Intensive Blood Pressure Management in the Post-Thrombolysis Setting: a real-world observational study

**DOI:** 10.1101/2023.04.19.23288841

**Authors:** Bethan Harper, Syrah Ranta, Harry McNaughton, Anna Ranta

**Affiliations:** Department of Neurology, Wellington Hospital; Department of Medicine, University of Otago, Wellington; Medical Research Institute of New Zealand, Wellington

**Keywords:** Stroke, blood pressure, thrombolysis, reperfusion, cerebral perfusion, intracerebral hemorrhage, outcomes

## Abstract

**Introduction:** Systolic Blood pressure (SBP) >180mmHg following stroke thrombolysis has been associated with increased bleeding and poorer outcome. Aiming for the guideline SBP of <180mmHg often leads to SBP overshoot as treatment is only triggered if this threshold is passed. We tested whether a lower target would result in fewer high SBP protocol violations.

**Methods:** This is a prospective, single centre, sequential comparison of two blood pressure protocols. Between 2013 and 2017, the guideline based post-thrombolysis SBP target of <180mmHg was compared with a new protocol aiming for 140-160mmHg. Primary outcome was rate of patients with SBPs >180 mmHg. Secondary outcomes included rate of SBP <120 mmHg, antihypertensive infusion use, symptomatic intracerebral haemorrhage (sICH), and 3-month functional independence (mRS 0-2). Results were adjusted for age, baseline function, and stroke severity using regression analysis.

**Results:** During the 23 months preceding and 18 months following the transition to the new protocol 68 and 100 patients were thrombolysed respectively. Baseline characteristics were similar between groups. The odds of one or more SBPs >180mmHg trended lower in the intensive group (adjusted odds ratio (aOR) 0.61; 95% CI 0.32-1.17; p=0.14). There was a higher rate of SBPs <120mmHg (aOR 3.09; 95% CI 1.49-6.40; p=0.002) in the intensive BP protocol group. sICH rate and 3-month mRS 0-2 were similar between groups.

**Conclusions:** The more intensive post-thrombolysis BP protocol led to a significant increase in sub-optimally low BP events with a non-significant trend toward fewer high BP protocol violations and unaffected patient outcomes.

## Introduction

Stroke remains one of the major causes of mortality and morbidity worldwide.^1^ Treatment with thrombolysis for acute ischaemic stroke (AIS) has improved outcomes, but carries an increased risk of symptomatic intracerebral haemorrhage (sICH).^2^ Up to 60% of patients presenting with AIS have hypertension.^3^ This may be attributable to a compensatory mechanism to increase cerebral perfusion pressure, pre-existing hypertension, pain, stress and inflammatory state.^4^ Systolic blood pressure (SBP) at presentation is an important prognostic factor, with both low and higher values associated with worse outcome (U-shaped curve). ^5,6^

The optimal target for blood pressure (BP) within the first 24 hours remains uncertain and is likely impacted by stroke type, cause of hypertension, type of reperfusion therapy received, if any, degree of recanalization, type and timing of the drug, BP variability, and speed of BP lowering.^2^ Some guidance is available specifically for the post-thrombolysis setting. The current American Heart Association/American Stroke Association and European Stroke Organisation guidelines recommend maintaining BP below 180/105mmHg during the first 24 hours post-thrombolysis.^7,8^ The ENCHANTED trial tested an intensive post-thrombolysis SBP target of 130-140 mmHg compared with the guideline target. There was no improvement in independence at 90 days but there was a significant reduction in any intracranial haemorrhage.^9^ In the Safe Implementation of Treatment in Stroke-International Stroke Thrombolysis Registry (SITS-ISTR) patients with SBP between 141-150mmHg had a four times lower risk of sICH than patients with SBP over 170 mmHg.^7^ Several observational studies have found that higher post-thrombolysis SBP and BP protocol violations have been associated with an increased risk of sICH, but also lower incidence of favourable functional outcomes.^10-12^ Taken together, these data suggest that SBP >180 mmHg are sub-optimal, for risk of sICH and possibly functional outcomes.

A review of our thrombolysis service found that post-thrombolysis SBPs of >180mmHg were not infrequent and that use of IV labetalol boluses as first-line management was associated with delays in achieving SBP control. If SBP of 180mmHg is the trigger for treatment then avoiding protocol violations of SBP >180mmHg is impossible as the protocol has to be violated in order for treatment to be initiated. We hypothesized that setting a slightly lower treatment trigger and treatment range would more consistently achieve SBP maintenance within guideline parameters without risking high rates of hypotension, and that use of protocolised continuous anti-hypertensive infusion may offer faster SBP target attainment and lower SBP variability.

The primary aim of this study was to assess whether a more intensive SBP management strategy in the first 24 hours post-thrombolysis using an ‘ideal range’ of 140-160mmHg, and a low threshold for initiation of IV antihypertensive infusion, would reduce the frequency of SBP > 180mmHg recordings.

## Methods

This is a prospective, single centre, open-label, unblinded observational study using a sequential comparison design to compare the rate of SBP > 180mmHg protocol violations with guideline based post-thrombolysis BP management compared to a more intensive strategy with an ‘ideal range’ of SBP 140-160mmHg and a low threshold for IV antihypertensive infusion.

At Wellington Regional Hospital, New Zealand the stroke service changed the protocol for management of hypertension after thrombolysis in mid-2014. The earlier protocol aimed for target SBP of <185mmHg pre- and <180mmHg post thrombolysis for the first 24 hours after treatment. The new protocol aimed for a target SBP of <185mmHg pre-thrombolysis and 140-160mmHg post-thrombolysis for the first 24 hours. Bolus IV Labetolol 10mg was to be used for SBPs above >185mmHg pre-thrombolysis. IV anti-hypertensive infusions were to be initiated if BP remained >160mmHg despite three or more IV Labetolol boluses in both the pre- and post-thrombolysis period. The protocol specified increments and decrements in the infusion rate depending on the SBP with frequent measurement and readjustment until the SBP was within range.

We identified patients from our prospectively collected thrombolysis database with supplementary retrospective chart review to collect additional baseline characteristics, BP recordings, and patient outcome data. Our patient group included all adult patients treated with IV thrombolysis for ischaemic stroke from January 2013 to January 2017. All patients had a clinical diagnosis of acute ischaemic stroke and all received thrombolysis within 4.5 hours of symptom onset. Computed tomography (CT) perfusion imaging was not in common usage during this period and a consistent thrombectomy service had not yet been implemented.

The primary outcome was number of patients experiencing one or more SBP of >180 mmHg during the first 24 hours following thrombolysis. Secondary outcomes included proportion of patients experiencing SBPs<160mmHg, <140 mmHg, <120 mmHg, or >200mmHg during first 24 hours, number of SBPs >180mmHg per patient, median SBP over 24 hours, >50% SBP drop between highest and lowest SBP recorded (to indicate variability), proportion receiving IV antihypertensives, sICH rate, and 3-month favourable modified Rankin Score (mRS) defined as 0-2 and also as mRS 0-1. sICH was defined as a National Institutes of Health Stroke Scale (NIHSS) deterioration of >4 points or death attributable to an ICH on post-tPA 24-hour CT imaging. All 24-hour CT images reporting any degree of bleeding were adjudicated by a blinded assessor (AR).

Sample size was estimated using a 60% rate of SBP >180mmHg for the standard protocol based on internal audit data and an estimated reduction of such events to 40% per patient with the intensive protocol. With a 95% confidence level and 90% study power this required a minimum sample size of 127.

Statistical analysis was performed using StataIC 17.0. Dichotomous and continuous variables were compared using chi square and either t-test for normal and Wilcoxen Rank-Sum for non-normally distributed continuous variables. Logistic regression incorporated common confounders and any differences in baseline characteristics of >0.1 using backward elimination technique to optimise model fit. Variables retained in the final model included age, baseline NIHSS, and pre-morbid mRS.

This study received no external funding and received Wellington Hospital institutional ethics approval under the category of ‘service audit.’ Individual patient consent was waived as the protocol change occurred for the primary purpose of improving guideline adherence as part of a service improvement project rather than for research purposes. Study data are available from the corresponding author upon reasonable request.

## Results

During the 23 months preceding and 18 months following the transition to the new protocol 68 and 100 patients respectively, with acute ischaemic stroke received IV thrombolysis. Baseline characteristics were similar between groups. (Table 1).

**Table 1:** Patient baseline characteristics by study group

| Patient Characteristic | Guideline<br>N=68 | Intensive<br>N=100 | P value |
| --- | --- | --- | --- |
| Age, mean (SD) | 72.8 (12.3) | 71.7 (15.6) | 0.65 |
| Ethnicity, n (%) |  |  | 0.43 |
| European | 50 (73.5) | 83 (83) |  |
| Maori | 5 (7.4) | 5 (5) |  |
| Indian | 5 (7.4) | 0 |  |
| Chinese | 1 (1.5) | 1 (1) |  |
| Pacific Island | 4 (5.8) | 7 (7) |  |
| Other | 1 (1.5) | 0 |  |
| Unknown | 2 (2.9) | 0 |  |
| Female sexr, n (%) | 25 (37) | 36 (36) | 0.92 |
| Hypertension | 40 (58.8) | 49 (49) | 0.21 |
| Diabetes | 14 (20.6) | 13 (13) | 0.19 |
| Atrial fibrillation | 18 (26.5) | 24 (24) | 0.72 |
| Anticoagulation | 5 (7.4) | 7 (7) | 0.93 |
| SBP pre thrombolysis,<br>median (range) | 159 (109-212) | 156 (102-241) | 0.79 |
| mRs prior to admission,<br>median (range) | 0 (0-4) | 0 (0-5) | 0.22 |
| NIHSS at presentation,<br>median (range) | 9 (2-30) | 9 (0-30) | 0.96 |

SD=standard deviation

Overall, the mean (95% Confidence Interval (CI)) SBP over the first 24 hours was 140.8 (137.8-143.9) in the intensive group and 147.1 (142.4-151.7) in the guideline group (mean difference (95% CI) 6.3 (0.97-11.6, p=0.02). Fewer patients in the intensive group had one or more SBPs >180mmHg (intensive 46 (46%) vs guideline 40 (59%)), but this was not statistically significant (adjusted Odds Ratio (aOR) 0.61; 95% CI 0.32-1.17; p=0.14). There was a statistically significant increase in rate of hypotension (SBP <120mmHg) recorded for the intensive management group (aOR 3.09; 95% CI 1.49-6.40; p=0.002). There was no difference in the number of patients with one or more SBP of >200, >160, <140 or <100 mmHg recorded or with a ≥50% difference between highest and lowest recorded SBP between groups. (Table 2).

**Table 2:**
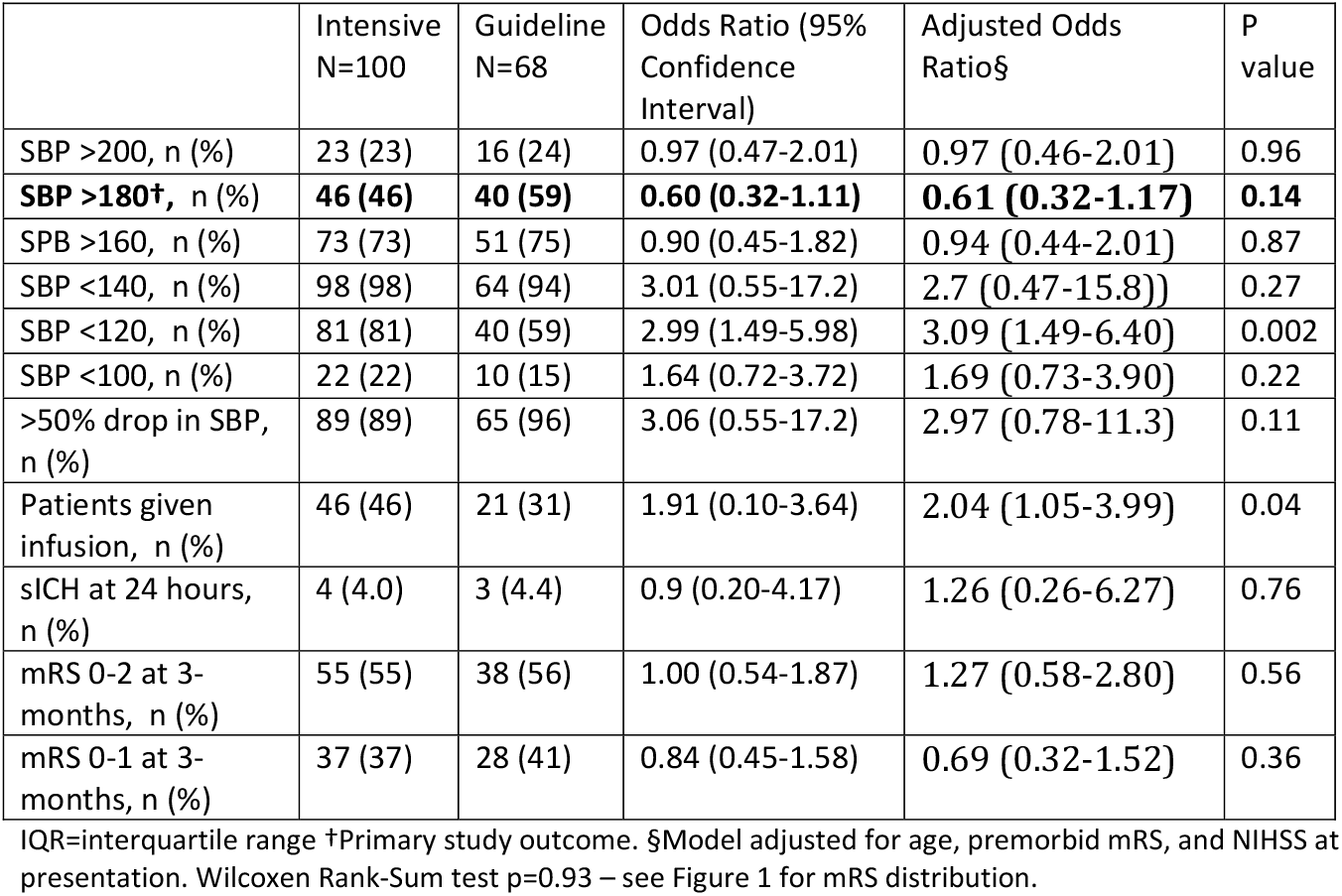
Blood pressures and patient outcomes by study group

Favourable outcomes (mRS 0-2) at 3 months and sICH were similar between groups with and without adjustment for potential confounders. More patients received an intravenous infusion of either Glyceryl Trinitrate (GTN) or Labetalol in the intensive BP protocol group (intensive group 46 (46%) vs guideline 21 (31%), aOR 2.04, (1.05-3.99) P=0.04). See Table 2 for additional detail.

We conducted additional exploratory analyses of number of SBPs >180 per patient, BP variability, and SBP at presentation. The mean number (SD) of SBPs >180 mmHg per patient was significantly lower in the intensive group (1.5 (0.22)) compared with 2.8 (0.49) in the guideline group; p=0.009). A similar pattern was observed for BPs >185mmHg: 0.84 (1.6) events per patient in the intensive and 1.8 (3.2) in the guideline group; p=0.002. For the study group as a whole, the number of high BP events was significantly correlated with poorer functional outcome (aOR=0.85 (0.73-0.99);p=0.038) and a higher rate of sICH (aOR-1.25 (1.06-1.48);p=0.01) adjusting for age, pre-morbid mRS and NIHSS at presentation. SBP at presentation and BP variability were not associated with outcome or sICH. (Details in Supplementary table).

## Discussion

Patients in the intensive group had a higher rate of intravenous antihypertensive use, lower mean SBP over the first 24 hours, non-significantly fewer SBP >180mmHg events, and significantly more SBP < 120 mmHg events. There was no difference in sICH rate or 3-month clinical outcome.

The lack of improved clinical outcomes is in keeping with the ENCHANTED trial,^9^ a phase 3 randomised control trial of intensive blood pressure lowering in the post-thrombolysis setting which pursued a more aggressive target (130-140 mmHg) than our protocol (140-160 mmHg) although resultant SBP levels were similar: ENCHANTED mean SBP intervention 138.8mmHg vs control 144.1mmHg at 1 hour and 144.3mmHg vs 149.8mHg respectively at 24 hours, compared with our mean 140.8 mmHg in the intensive group vs 147.1 mmHg in the guideline group over the 24 hour period. Similar to our results ENCHANTED failed to demonstrate an improvement in either 3-month mRS or sICH rate although they did find a reduction in any ICH.

Our study was powered to detect a difference of 20% in high BP events between the groups and we observed a reduction of 15%, arguably still clinically significant but requiring a larger study to demonstrate statistical significance. The higher frequency of very low SBPs cannot be ignored. One reason for this may have been too much attention to SBP at the higher end of the scale so that nurses were less attentive when the SBP was in the ‘ideal range’ but falling, and delayed reduction and/or stopping of the antihypertensive infusion. The protocol for changing the infusion rate may have erred on the side of too aggressive lowering down to a too low floor level (ie SBP=140). If so, these issues could be remedied by training and a slightly higher floor to the ‘ideal range’ – eg SBP=150. We acknowledge, along with others, that BP management post-thrombolysis needs to be individualised taking into account stroke type, presence of large vessel occlusion, success of recanalization, and other factors.^2^ For example, it is likely that sICH risk is highest in recanalized larger strokes (implying tighter SBP control is required) while infarct growth due to hypoperfusion is of greatest concern in large vessel occlusion patients who did not recanalize where somewhat higher SBP targets may be appropriate.

The choice of antihypertensives and rapidity of BP lowering may be relevant to successful outcomes. We note that the INTERACT 4 trial is testing very early ambulance-based BP lowering in acute ischaemic stroke or ICH is using the antihypertensive agent Urapidil -an α_1_-adrenoceptor antagonist and a 5-HT1A receptor agonist – which may have advantages over labetalol and GTN.^13^

This study had several limitations. The relatively small sample size may have introduced type 2 error, and was underpowered for the difference in SBP detected between the groups.

The observational sequential design carries the usual risks of potential confounding. The single centre nature may limit generalisability. Finally, patients with less well controlled BP and on IV infusions had more BPs recorded than those with primarily normal range BPs which may have led to potential reporting bias especially as regards BP extremes. Strengths of the study were the prospective acquisition of data, ‘real-world’ comprehensive coverage and completeness of follow-up.

A more aggressive approach to early BP lowering requires higher use of IV antihypertensive medication -in ENCHANTED, 63% of intensive patients vs 35% of control patients received intravenous medication, while in our study 46% of intensive vs 31% of guideline patients received intravenous medication. This has implications for nursing resource, cost of medications and equipment and the potential for IV site related complications.

At this stage, the absence of clear benefit in our study (and ENCHANTED) and evidence of potential for harm argue against a more aggressive approach. We are trialling a new protocol with an ‘ideal range’ of SBP 150-170 combined with training to prevent low SBP events and a more tailored approach to patients post-thrombectomy based on the presence or absence of successful recanalization.

## Data Availability

Study data are available from the corresponding author upon reasonable request.

## Abbreviations

(s)ICH: (symptomatic) intracerebral hemorrhage
(S)BP: (systolic) blood pressure
mRS: modified Rankin Score
NIHSS: National Institute of Health Stroke Scale
CT: Computed tomography

## Acknowledgements

None

## Disclosures

None

**Figure 1:**
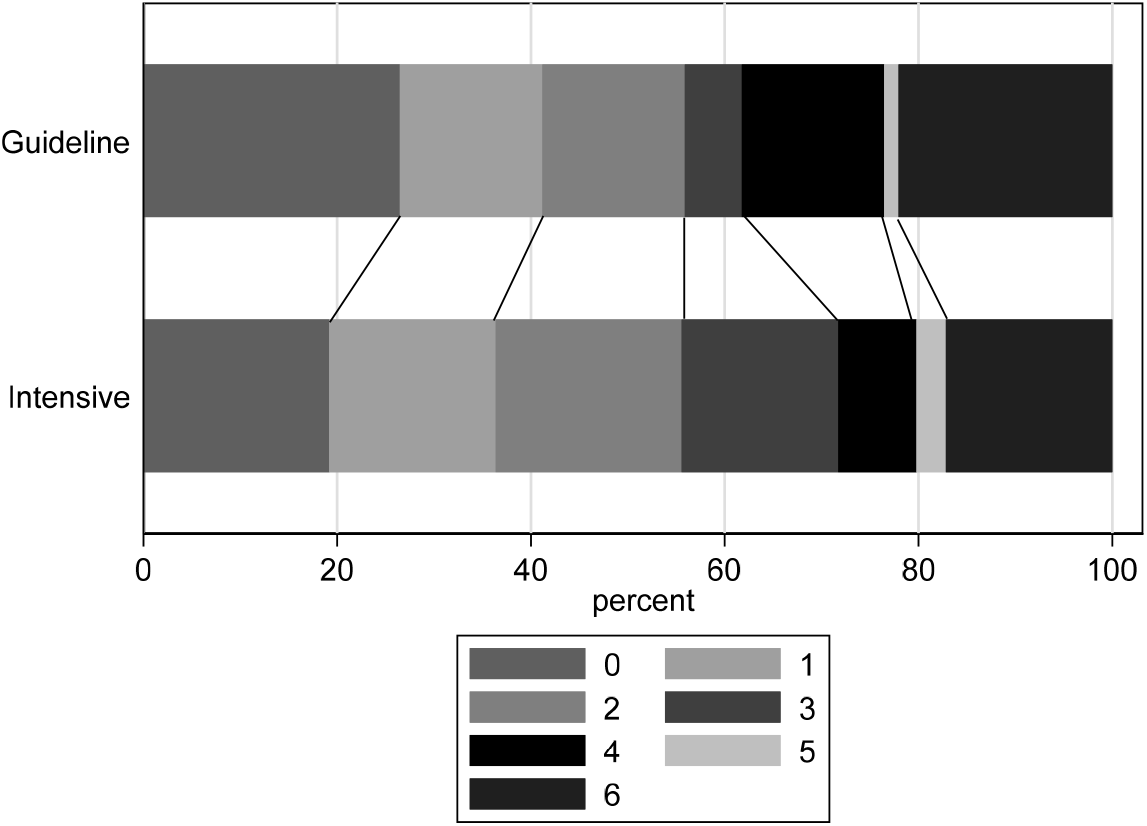
3-month modified Rankin Score Grotta Chart (median (IQR) intensive: 2 (1-4), guideline: 2 (0-4); Wilcoxen Rank-Sum p=0.93)

